# HIV Pre-Exposure Prophylaxis Scale-Up in Mozambique from October 2021 to December 2023

**DOI:** 10.1101/2024.11.25.24317762

**Authors:** Gonçalves Maibaze, Nuno Gaspar, Jessica Seleme, Meghan Swor, Aleny Couto, Inacio Malimane, Marcos Canda, Ana Paula Simbine, Mark Troger, Evangelina Namburete

## Abstract

In 2021, The Mozambique Ministry of Health began offering HIV pre-exposure prophylaxis (PrEP) in all provinces. The number of new clients increased more than three-fold within two years. Among 51,848 new people receiving PrEP by December 2023, the largest groups were females and adolescent young adults.

## Introduction

Robust combination prevention strategies are crucial to ending AIDS as a public health threat by 2030. Aiming to minimize the number of new infections, HIV pre-exposure prophylaxis (PrEP) is a core component of combination prevention recommended by the World Health Organization (WHO).^1-3^ PrEP lowers the risk of HIV acquisition among HIV-negative people by using antiretroviral medication.^1,2^ Several options are available, including daily oral and injectable long-acting PrEP. In general, WHO recommends that healthcare providers prescribe PrEP to individuals who could benefit from PrEP, for example: a) Individuals who request PrEP; OR b) Individuals with likely ongoing HIV exposure, which may include any of the following:

i a sexual partner living with HIV who is not virally suppressed on HIV treatment;
ii recent or probable future inconsistent use of condoms for vaginal or anal sex;
iii a recent sexually transmitted infection (STI);
iv recent PEP use for sexual exposure to HIV, especially for individuals who have used PEP more than once.^2^

In 2017, Mozambique piloted PrEP programming in three provinces (Nampula, Zambezia, and Sofala), initially prioritizing key populations (KPs), serodiscordant couples, adolescent girls, and young women (AGYW), and pregnant and breastfeeding women (PBFW). Given high acceptance among people utilizing PrEP, Mozambique scaled up PrEP nationally in 2021 to all at-risk individuals who met eligibility criteria: a person with an HIV-negative test within the last 3 months with no suspicion of acute HIV infection based on screening signs and symptoms, without contraindications to PrEP drugs, and with signed informed consent.^4^

To understand progress in scaling–up PrEP in Mozambique, we analyzed program data from 2021 to 2023.

## Methods

We analyzed U.S. President’s Emergency Plan for AIDS Relief (PEPFAR) Monitoring, Evaluation, and Reporting quarterly data from October 2021 to December 2023 from all PEPFAR-supported sites in Mozambique.^1^

A “new PrEP client” was defined as a person who is HIV-negative and newly enrolled in PrEP services. “Continuity on PrEP” was defined as a PrEP client who returned for a PrEP follow-up visit or re-initiation, excluding people newly enrolled ^4,5^ An “HIV seroconversion” was defined as a PrEP client who tested positive for HIV using the Mozambique national algorithm after ≥3 months of PrEP enrolment.^4^ Following Mozambique Ministry of Health PrEP guidelines, high-risk groups were PrEP clients aged 15–24 years and KPs.^4^ Data were analyzed by quarter, sex, province, and high-risk groups.

## Results

From October 2021 to December 2023, new PrEP clients increased from 15,733 to 51,848 (females = 9,575 [60.8%] to 33,949 [65.5%]; 15–24 years = 7,321 [46.5%] to 26,786 [51.7%]; KPs = 3,646 [23.2%] to 12,613 [24.3%]). Overall provincial growth rate ranged from Manica Province, with a growth rate of 26% (from 4,053 to 5,115), to Maputo City, with a growth rate of 3,405% (from 95 to 3,330) (Table 1).

**Table 1.** Number of New PrEP Clients, by Province of Mozambique, and Percentage Increase between October–December 2021 to October–December 2023.

| Provinces | Number of New PrEP Clients |  | % increase |
| --- | --- | --- | --- |
|  | Oct-Dec 2021 | Oct-Dec 2023 |  |
| Military Mozambique | 819 | 1,254 | 53% |
| Cabo Delgado | 106 | 1,631 | 1,439% |
| Maputo City | 95 | 3,330 | 3,405% |
| Gaza | 76 | 2,065 | 2,617% |
| Inhambane | 67 | 1,589 | 2,272% |
| Manica | 4,053 | 5,115 | 26% |
| Maputo | 479 | 1,837 | 284% |
| Nampula | 5,494 | 8,907 | 62% |
| Niassa | 108 | 1,717 | 1,490% |
| Sofala | 256 | 2,206 | 762% |
| Tete | 148 | 3,604 | 2,323% |
| Zambezia | 4,032 | 18,593 | 361% |
| **National** | **15,733** | **51,828** | **229%** |

During the October–December 2021 quarter, a total of 4,920 clients returned for PrEP follow-up or a re-initiation visit (females = 3,068 [62.4%]; 15–24 years = 1,974 [40.1%]; KPs = 1,025 [21.0%]). Between October–December 2023, a total of 22,273 clients returned for PrEP follow-up or a re-initiation visit (females = 14,153 [63.5%]; 15–24 years = 10,029 [45.0%]; KPs = 3,639 [16.3%]) (Table 2).

**Table 2.** Number of PrEP Clients who returned for continuity or re-initiation of PrEP, by Province of Mozambique, and Percentage Increase between October–December 2021 to October–December 2023.

| Provinces | Number of PrEP Continuity Clients |  | % increase |
| --- | --- | --- | --- |
|  | Oct-Dec 2021 | Oct-Dec 2023 |  |
| Military Mozambique | 399 | 323 | -19% |
| Cabo Delgado | 2 | 616 | 30700% |
| Maputo City | 1 | 482 | 48100% |
| Gaza | 36 | 749 | 1981% |
| Inhambane | 67 | 642 | 858% |
| Manica | 813 | 2152 | 165% |
| Maputo | 1 | 1356 | 135500% |
| Nampula | 1759 | 8224 | 368% |
| Niassa | 0 | 261 | 26100% |
| Sofala | 4 | 494 | 12250% |
| Tete | 4 | 563 | 13975% |
| Zambezia | 1834 | 6411 | 250% |
| **National** | **4,920** | **22,273** | **353%** |

## Discussion

In 2 years, new PrEP clients increased by 230%, with the largest groups receiving services being females, adolescents, young adults, and KP, who are at high risk for acquiring HIV infection. Continued scale-up could be supported by improved demand creation, community education, patient literacy, and further support for the continuity of PrEP for those in need.

While the scale-up has been impressive, the literature supports wider dissemination of PrEP literacy to both providers and clients to increase equitable PrEP uptake among targeted at-risk populations as well as improve the use of PrEP drugs as prescribed.^6-8^

Our findings support further expansions of our PrEP program to reach more people, including developing a communication package for literacy, which will include a PrEP communication campaign, quality improvement interventions, a demand creation strategy, and an assessment of PrEP acceptability.

There are several limitations of the analysis. Analyzed programmatic data quality may be impacted by reporting challenges. However, continual data quality improvement activities minimized challenges. Further, reported data did not capture contextual differences, which could be useful for understanding percentage increases. Finally, clients could seek services at any site, regardless of residence. Thus, some people might have been counted as being on PrEP more than once.

## Data Availability

All data produced are available from PEPFAR Monitoring, Evaluation, and Reporting program data.

https://data.pepfar.gov/datasets#PDD

## Acknowledgements

We would like to extend our gratitude to the U.S. Centers for Disease Control and Prevention (CDC), the U.S. Agency for International Development (USAID), and the Ministério da Saúde (MISAU) of Mozambique for their support and collaboration in this manuscript.

We are grateful to the healthcare providers, clinicians, and staff who have dedicated their time and effort to the implementation and expansion of the HIV pre-exposure prophylaxis (PrEP) program across Mozambique. Their unwavering commitment has been instrumental in reaching this coverage of PrEP services.

## Funding Acknowledgement

This manuscript has been supported by the President’s Emergency Plan for AIDS Relief (PEPFAR), through the Centers for Disease Control and Prevention (CDC), The findings and conclusions in this manuscript are those of the author(s) and do not necessarily represent the official position of the funding agencies.

## Footnotes

1 This activity was reviewed by CDC, deemed not research, and was conducted consistent with applicable federal law and CDC policy. See e.g., 45 C.F.R. part 46.102(I)(2), 21 C.F.R. part 56; 42 U.S.C. 241(d); 5 U.S.C. 552a; 44 U.S.C. 3501 et seq.

